# *“Men’s is the only and final word*.” A qualitative study of community midwives’ insights on violence against women and girls in Yemen, including the forms, causes, and responses of survivors and their families

**DOI:** 10.1101/2025.01.02.25319891

**Authors:** Marwah Al Zumair, Hussein Zaid, Luz Marina Leegstra, Raisa Ferrer Pizarro, Priya Shreedhar, Monia Al Zumair, Lamya Bawahda, Albrecht Jann, Lauren Maxwell

## Abstract

Violence against women and girls (VAWG) is a serious human rights violation that has intensified in Yemen due to war, displacement, and pandemics. The cultural stigma surrounding discussions of VAWG, particularly sexual violence, compounded by a lack of resources for affected individuals, poses significant challenges for research and program implementation. This study involved qualitative interviews with 20 community midwives from the National Yemeni Midwifery Association across four governorates to explore the causes, forms, and consequences of VAWG and identify potential resources for survivors. A Yemeni physician trained in ethical VAWG research conducted the interviews, which were analysed thematically by two researchers. The findings indicated that physical partner violence was widely recognised as a crime; however, women and girls facing sexual violence often faced severe social ramifications, including ostracism or violence aimed at preserving family honour, leading to underreporting. Midwives identified patriarchal culture and the ongoing economic crisis as key contributors to VAWG. Health-related consequences for survivors included physical injuries such as vaginal tears and obstetric fistula. Additionally, midwives reported that survivors encountered humiliation within the healthcare system. Most women and girls did not seek help; however, when they did, they turned to their families or local leaders for support. To effectively understand, prevent, and address VAWG in Yemen, strategies should prioritise the safety and needs of women and align with cultural values. Insights from community midwives can guide the development of VAWG-focused community-led initiatives.

## Introduction

Violence against women and girls (VAWG) is a violation of human rights. Addressing VAWG is central to achieving several of the Sustainable Development Goals (SDGs), particularly SDGs, which include eliminating violence against women. Globally, an estimated one in four women ever experiences intimate partner violence (IPV), the most prevalent form of VAWG [1]. VAWG increases during displacement, conflict, economic crisis, and pandemics, all of which have affected Yemen in recent years [2–4]. This multifaceted crisis has disproportionately affected women and girls who are often left as the sole providers for their families because of the absence or incapacitation of male family members due to the ongoing conflict or job loss due to the ongoing war and recent COVID-19 pandemic [4,5]. The few studies that have measured VAWG in Yemen have found that interpersonal violence is highly prevalent [6,7]. The 2013 Yemeni Demographic and Health Survey (DHS), the only DHS conducted in Yemen since 1992 and the only one to ask violence-related questions, found that 92% of women and girls aged 15–49 had witnessed DV in their homes [8]. The 2013 DHS estimated the national prevalence of female genital mutilation (FGM) at 19%, with significant regional variation [8]. The prevalence ranged from 0% in Al-Baidha to 85% in Al-Mhrah [8]. A country brief conducted by UNICEF also found that the median age of marriage varied across Yemeni governorates. The median age was lower in rural and higher in urban regions (17.9 versus 18.9) and the lowest among girls living in Al-Jawf (17 years) [9]. These findings suggest the need to explore geographical heterogeneity in VAWG and factors relating to exposure.

The United Nations Development Programme (UNDP) ranked Yemen in the last decile of its 2015 Gender Inequality Index (168 out of 188 countries) [10]. Societal VAWG, defined as practices, norms, and beliefs that prevent women and girls from exercising their full human rights, takes many forms in Yemen [11]. Yemeni women and girls often eat the food left over after their male counterparts have eaten, and most families preferentially educate and provide medical care to their male children. The 2013 DHS found that only 43% of Yemeni women and girls over six years of age had ever attended school, and 47% of women and girls were illiterate [8]. Laws related to inheritance, custody, infidelity, divorce, and legal testimony favour men over women [11]. A 2010 UN country assessment on gender-based violence (GBV) described several forms of violence against women which may differ from other geographies. For instance, blood money, known as “Diya,” the financial compensation paid to the victim’s heirs as a form of punishment to the killer for the death of a spouse or relative for a woman is half of that for a man [12]. The report also found that Ethiopian and Somali migrants to Yemen faced an increased risk of sexual violence [12]. The 2013 DHS found that half of women and girls aged 15–49 believed that a husband is justified in beating his wife [8].

The ongoing conflict and the COVID-19 pandemic have increased the spectrum, severity, and incidence of VAWG in Yemen [6]. Women and girls whose husbands or fathers were killed in the ongoing conflict face increased violence through child marriage, forced and exchange marriage, sex trafficking, and increased instances of harassment and rape [7,13]. Although the 2013 DHS reported that the median age of marriage was eighteen [8], smaller-scale studies found a dramatic increase in child marriage after the nationwide socioeconomic disruption caused by the 2014 war, as families that were no longer able to feed their children saw child marriage as the only way to feed their children [9]. A recent assessment across northern and southern Yemen reported that 72% of the interviewed adolescent boys and girls were married at 13–15 years and 17% at 8–12 years old [7]. Travel or tourist marriages where Yemeni girls are “married” to a man from an Arabian Gulf country and the groom annuls the marriage several days to months later when he finishes his vacation are a novel form of VAWG that has emerged from the longstanding economic crisis [14]. While the male spouse can easily dissolve the “tourist marriage,” women and girls confront different forms of stigma and adverse physical and psychological consequences and remain legally bound to the tourist marriage, requiring them to take a number of steps to dissolve the marriage [14].

While VAWG has increased in Yemen, the mechanisms available for women and girls to avoid, mitigate, confront, or address this violence have become more limited [15]. Given the lack of information on VAWG and related services in Yemen, we conducted a qualitative interview study with members of the National Yemeni Midwifery Association (NYMA) to understand the underlying causes of, forms, and consequences of VAWG and identify resources that can support women and girls who experience violence. NYMA has existed in Yemen since the 1990s and operates in every Yemeni governorate. NYMA midwives are trusted health providers who live and work in the community and are often the only point of conduct between women and girls and the formal health care system, especially in rural communities. Conducting this formative research with community members and developing culturally relevant safety planning materials are essential precursors for the safe and ethical conduct of VAWG-related research with survivors. We conducted this formative research to help ensure that future violence-related research and interventions in Yemen are grounded in the local context and responsive to deeply ingrained societal norms and practices.

## Methods

We conducted one-time, in-depth interviews (IDIs) with 20 NYMA midwives between 1 November and 31 December 2021. The community midwives were based in urban and rural areas in four Yemeni governorates. Aside from five interviews conducted over the phone because midwives were located in regions that were difficult to access, interviews were conducted in person. The first author, a Yemeni physician trained in conducting qualitative research as well as the ethical conduct of VAWG-related research, led the interviews in the Yemeni dialect of Arabic, paying attention to non-verbal cues such as the midwifes’ facial expressions and recording debriefing notes after each interview. The interviews followed a semi-structured interview guide prepared by researchers and clinicians with experience in the VAWG field and Yemen. The interview guide was composed of questions and probes that reflected the study’s objectives, which included identifying what the midwives knew about the spectrum of violence, perpetrators, and consequences; midwives’ experience working with women and girls who experienced violence; their and the community’s response to VAWG; and barriers and facilitators to serving women and girls who experience violence, including access to shelters and legal services for survivors. The interview guide and protocol are available on the Open Science Foundation (https://doi.org/10.17605/OSF.IO/F7QSJ). The interview guide was piloted with three participants and refined throughout the interviews to address emerging themes. Following the pilot, several redundancies in the guide were removed. The midwives provided verbal consent for the one-time, hour-long interview. The interviews were conducted until saturation was reached, defined as no new themes emerging [16].

We conducted IDIs with community midwives rather than with Yemeni women and children for several reasons. First, Yemeni community midwives are trusted frontline healthcare providers. They are often the first and only place where women and girls who experience violence interact with the health system, particularly in rural areas [17]. Their close relationships with women, cultural sensitivity, and direct exposure to cases of GBV enable them to provide unique insights into the context that shapes the experience and response of women and girls who experience violence and healthcare response to VAWG [18]. Secondly, by asking midwives about VAWG that they have learned about through their work, we reduced the risk of retaliation or harm for survivors who participate in VAWG-related research in a setting where stigma and honour-based violence are prevalent and asking questions about women or girls’ direct experience of violence may put them at risk. Lastly, before the initiation of any research with VAWG survivors, the research team must have an excellent understanding of the context for violence and reporting and, have co-developed safety planning materials and identified an accessible support network for women and girls who experience violence. Our formative interviews with community midwives provide an essential foundation for developing safe, ethical future research that is responsive to the challenges of measuring and addressing VAWG in the Yemeni context.

### Identification and recruitment of study participants

Participants were recruited in collaboration with the NYMA’s manager, who discussed the study with NYMA midwives and provided a contact list of midwives willing to participate to the study manager. As NYMA has branches across Yemen, we selected midwives from different regions to explore heterogeneity in their responses. We contacted the first potential participant on the list in that governorate and moved to the next participant if we could not reach the first one. No midwives that we reached to discuss the study refused participation. Midwives were contacted a week before the interview to discuss the interview format and objectives, and the interview was scheduled based on their availability. The interviews were conducted either at the NYMA office or at a location agreed upon by the researcher and the midwife. The selection process ensured midwives were from different governorates with various geographical features, such as highlands, agricultural areas, and coastal regions, and lived in different urban, semi-urban, and rural settings. We also selected midwives from various levels of healthcare, such as primary health facilities, private clinics, hospitals, and governorate or district health offices, to explore heterogeneity across these levels.

### Researcher characteristics and reflexivity

The first author, a Yemeni physician with experience in qualitative research and applied work in sexual and reproductive health and rights (SRHR) in Yemen, conducted the interviews. All authors were trained in the ethical conduct of VAWG-related research, and several have applied experience in community-based participatory research. Four authors were Yemeni physicians or health care providers who have worked in Yemen, and a fifth author had lived and worked in Yemen.

### Ethical considerations

Ethical approval of the study protocol was obtained from the Universitatsklinikum Heidelberg Research Ethics Committee (Approval No. S-345/2021) and the Sana’a University of Medical Sciences (Approval No. 1548). Participants provided informed verbal consent because the COVID-19 pandemic was ongoing at the time of data collection and because the interview did not ask for participants’ data or their experience of VAWG. While midwives reported their experiences while working in SRH, we followed the best practice for research with survivors of violence [19]. Interviewers were trained in conducting VAWG-related research, and we ensured participants’ privacy during the interview. Because of the limited financial support for the project, we were not able to provide psychological support to the respondents who shared their experiences of witnessing or supporting survivors. Further ethics considerations are reported in the PLOS Inclusivity in Global Research Questionnaire (S1 Text).

### Data analysis

The interview recordings were transcribed verbatim in Arabic and translated into English. Identifying information, like patient names, was removed from the written transcripts. The research team met 1-2 times weekly during the interview and analysis period. Three researchers used MAXQDA to independently conduct a thematic analysis of the English transcripts, employing inductive and deductive coding techniques. The research team created an initial codebook based on the interview guide and a literature review. The literature review search strategy and codebook are presented in Appendix Tables S1 and S2. Initial codes were developed and gradually transformed into more focused codes and, eventually, conceptual categories. Repeated reading and comparison of the coded data ensured reliability across the application of the codes, which was conducted independently by the two researchers. Themes and corresponding quotations in the original Arabic and their English-language translation are provided in Appendix Table S3. Findings are presented in keeping with the COREQ (COnsolidated criteria for REporting Qualitative research) Checklist for reporting qualitative research, provided as Appendix Table S4 [20,21].

## Results

### Study population

Demographic information on the midwives and their training and experience in VAWG are presented in Table 1. We conducted 20 in-depth interviews with NYMA community midwives in four Yemeni governorates. Close to two-thirds of the interviews were conducted face-to-face, while one-third took place over the phone due to limited access to the regions where the midwives stayed. Half of the midwives (N=10) were between 30 and 39 years old, 35% (N=7) were between 40 and 49 years old, and 15% (N=3) were over 50 years old. Most midwives (70%; N=14) were married, 20% (N=4) were single, 5% (N=1) were divorced, and 5% (N=1) were widowed. More than half of the midwives (63%; N=11) had between one and five children, 19% (N=3) had no children, and another 19% (N=3) had more than five children. The selection of midwives was distributed among the included governorates. Sana’a Amanat al-Asimah was the place of origin for 10% (N=2) of the midwives, Sana’a governorate 25% (N=5), Taiz 35% (N=7), Aden 25% (N=5), and Ibb 5% (N=1). Five midwives (25%) worked in each of the governorates of Sana’a, Sana’a Amanat al-Asimah, Taiz, and Aden. Almost half (55%; N=11) of the midwives had intermediate diplomas and were classified by the high medical council as community midwives. Another 20% (N=4) of the midwives possessed a high diploma in midwifery and were classified as professional midwives. Another 25% (N=5) of the interviewees were midwifery trainers. All interviewed midwives had over ten years of experience in their profession. Close to half (45%; N=9) of the midwives worked at hospitals, 30% (N=6) at health facilities, 15% (N=3) at district, governorate health offices or the Ministry of Health, 5% (N=2) for NYMA, and 5% (N=1) in private clinics. Geographically, 60% (N=12) lived and practised in urban areas and 40% (N=8) in rural areas. A few midwives had received SRHR-related training, including on the minimal initial service package (MISP) [22], empowering women, SRHR, and identifying and clinically managing violence survivors. However, in the interviews, midwives reported that VAWG-related training had been significantly reduced since the war. VAWG, women’s SRHR, and women’s empowerment were deemed sensitive, and midwives found that the new government was resistant to supporting or investing in initiatives that were not socially acceptable.

**Table 1.** Demographic characteristics of interview participants (N=20)

|  | N | % |
| --- | --- | --- |
| **The interview conducted** |  |  |
| Face to-Face | 15 | 75% |
| Over the phone | 5 | 15% |
| **Age (years)** |  |  |
| 30-39 | 10 | 50% |
| 40-49 | 7 | 35% |
| 50 or older | 3 | 15% |
| **Marital status** |  |  |
| Single | 4 | 20% |
| Married | 14 | 70% |
| Widow | 1 | 5% |
| Divorced | 1 | 5% |
| **Parity** |  |  |
| No children | 3 | 19% |
| 1-5 Children | 10 | 63% |
| > 5 children | 3 | 19% |
| **Governorate of origin** |  |  |
| Sana'a Amanat al-Asimah | 2 | 10% |
| Sana'a Governorate | 5 | 25% |
| Taiz | 7 | 35% |
| Aden | 5 | 25% |
| Ibb | 1 | 5% |
| **Governorate where midwives work** |  |  |
| Sana'a Amanat al-Asimah | 5 | 25% |
| Sana'a Governorate | 5 | 25% |
| Taiz | 5 | 25% |
| Aden | 5 | 25% |
| **Profession** |  |  |
| Community midwife (intermediate diploma) | 11 | 55% |
| Professional Midwife (high diploma) | 4 | 20% |
| Midwife trainer | 5 | 25% |
| **Number of years in the profession** |  |  |
| Up to 10 years | 0 |  |
| Over ten years | 20 | 100% |
| **Where the midwives work** |  |  |
| Hospitals | 9 | 45% |
| Health facility | 6 | 30% |
| GHO/DHO | 3 | 15% |
| NYMA | 1 | 5% |
| Private clinic | 1 | 5% |
| **Population served by the midwife** |  |  |
| Urban | 12 | 60% |
| Rural | 8 | 40% |

DHO=District Health Office; GHO=governorate health office; NYMA=National Yemeni Midwife Association

### Emotional violence

Midwives reported that the verbal violence they heard about from women and girls took many forms, including shouting, mistreatment, favouritism toward male children, insults, and humiliation. Midwives described emotional violence as confining girls at home, threats of divorce, and threats to abduct or harm children. Women and girls reported to midwives that emotional violence led to psychological harm, social isolation, and challenges in forming and maintaining healthy relationships.

> *“Verbal violence involves a man’s negative behavior towards his wife and a lack understanding. His language often includes aggressive words and can sometimes escalate to physical violence.” (MW-02, Amanat al-Asimah, urban, age 25-34)*

### Physical violence

Women and girls shared their experiences of physical violence with midwives and sought their help for support.

> *“Once, a runaway girl came to me, wanted me to clean her wounds resulting from her husband’s beating. She was in a terrible situation.” (MW-03, Amanat al-Asimah, urban, age 35 or over)*

Midwives indicated some other examples of physical violence, more common in rural settings, including heavy labour and the use of pesticides by pregnant women.

> *“Violence can be from the parents or a husband. Physical violence is not limited to beatings, but also through assigning women hard work such as farming and the use of pesticides during pregnancy.” (MW-07, Amanat al-Asimah, urban, age 35 or over)*

According to midwives, domestic violence was the most common form of VAWG.

> *“Domestic violence is more prevalent because the husband thinks that nobody knows about their conflicts at the house unless the woman speaks.” (MW-01, Amanat al-Asimah, urban, age 35 or over)*

### Sexual violence

The majority of midwives recounted stories of women and young girls being raped, either by family members or strangers, sometimes resulting in pregnancy. They noted that the escalation of conflict in Yemen has heightened the risk of women facing kidnapping, rape, sexual harassment, and even sex trafficking, as families are increasingly driven to desperate measures for survival.

> *“I know a girl who was an orphan on her mother’s side, and her father pressured her to do anything to make money. Eventually, she resorted to sex work and contracted AIDS, leading her to visit the AIDS centre.” (MW-09, Sana’a Gov, rural, age 25-34)*

Women and girls reported to midwives that they had experienced marital rape, which is not recognised as a crime in Yemen. Sexual violence resulted in severe physical consequences, including vaginal tears and heavy bleeding, particularly among girls.

> *“As midwives, we see cases of sexual abuse by husbands. For example, I saw some cases complaining of violence from their husbands during sex (forced or violent sex, which also includes beating). So, they come to us because of bleeding or tears, etc. I think this is abnormal behaviour.” (MW-16, Aden, urban, age 35 or over)*

Midwives also reported rape by other family members or strangers.

> *“The mother had a disabled child whom she regularly took to the…Hospital…for physiotherapy. So, her daughter was at home alone. While the daughter was collecting prayer clothes in the yard, someone raped her. I gave her eight contraceptive pills because I was afraid the girl would get pregnant.” (MW-03, Amanat al-Asimah, urban, age 35 or over)*

Midwives reported that Yemeni society stigmatises women and girls who experience sexual assault and frequently posits them as the reason for the sexual assault, which complicates reporting. Midwives reported that survivors of sexual violence are frequently forced to marry their rapists or are sometimes accused of immoral acts and imprisoned.

> *“We have a criminal police unit; If cases of sexual violence are identified or if there’s suspicion of pregnancy in an unmarried woman, they detain her.” (MW-15, Taiz, urban, age 35 or over)*

In addition to social stigma, midwives reported that inadequate legal protection, lack of trust in law enforcement and the legal system, generalised violence (e.g., the potential for experiencing violence in transit to report violence), and the fact that sexual violence, including sexual harassment, is only recognised if the woman’s hymen is ruptured, itself highly stigmatised, led to underreporting of experiences of sexual violence.

> *“Yemen’s laws on sexual violence are weak, almost non-existent. Even if a woman speaks up, she often cannot seek proper justice unless there is evidence of hymen penetration.” (MW-02, Sana’a Amanat al-Asimah, urban, age 25-34)*

### Early, forced and tourist marriages

Midwives cited the prolonged conflict and the associated displacement, insecurity, and poverty as having increased the prevalence of early marriage above pre-war levels.

> *“Some families are convinced to marry off their daughters at an early age. Some people believe a girl should marry in these current economic situations. In my neighborhood, I know a father who married off his young daughter while she was in the seventh grade. I think some people do this to get rid of the burden of girls.” (MW-16, Aden, Urban, age 35 or over)*

> *“Society marries off girls at an early age to get rid of their responsibilities.” (MW-08, Sana’a Gov, rural, 35 years or over)*

> *“I cannot intervene in issues of early marriage because the father believes he owns the girl and can do whatever he wants; he considers it a family matter.” (MW-03, Amanat al-Asimah, urban, aged 35 or over)*

Midwives suggested that the financial hardship faced by many Yemeni families was the driving force behind the emergence of tourist marriage.

> *“A law must be made to prevent the marriage of girls under the age of 18 because the situation has escalated so much. If a Gulf person comes and has money, he will sell his daughter even if she is ten years old.” (MW-14, Taiz, rural, age 25-34)*

In addition to early marriage being a form of violence, midwives reported that families resorted to physical violence to force girls into marriage.

> *“Brothers beat their sisters to force them into marriage, just as fathers do, who physically abuse them in order to marry them off (early child marriage).” (MW-03, Amanat al-Asimah, urban, age 35 or over)*

Midwives reported that forced early marriage had, in addition to physical consequences, severe mental health consequences for girls, including suicide to escape their situation.

> *“A 13-year-old girl drank poison because she was forced to marry her cousin.” (MW-01, Amanat al-Asimah, urban, age 35 or over)*

### Family honour-related violence

Midwives raised concerns about the practice of virginity testing conducted on women before marriage, a common practice associated with maintaining family honour. Midwives reported that if a husband did not observe his wife bleeding following sexual intercourse on the first night of the marriage, they would request virginity testing to verify the woman’s virtue.

> *“A woman usually comes to us with her husband, mother, or father. Most of the cases who come to the hospital for a virginity test either have a history of rape or a bride who wants a report to prove their virginity.” (MW-02, Amanat al-Asimah, urban, age 25-34)*

When a woman or girl’s virginity is questioned, the woman or girl is at risk of being divorced and even murdered under the guise of preserving family honour.

> *“Once, a case came to me on her wedding night. The bride’s and husband’s families were all armed. They wanted me to let the bride’s mother enter the examination room with us. I was terrified by this situation and refused to expose her and say what I saw.” (MW-07, Amanat al-Asimah, urban, age 35 or over)*

### Gendered inequities in access to food, medical care, education, and legal resources

Midwives described how gender inequitable societal norms were a form of violence and also exacerbated other forms of violence by preventing women from leaving abusive relationships, help-seeking, or reporting. Midwives reported that women and girls had limited mobility.

> *“Men are always given priority; men’s is the only and final word. Men believe that a woman does not have the right to discuss or speak; she should only say ‘yes.’” (MW-20, Aden, urban, age 35 or over)*

In cases of separation, midwives found that women were required to seek legal assistance to obtain what is known as “Khul,” a form of annulment recognised by the court. In contrast, men can unilaterally declare “Talak” to divorce their wives without resorting to legal intervention. Midwives reported that women and girls told them about their own or their family and friends’ experiences of being divorced for difficult-to-justify reasons or even to marry another woman.

> *“In the divorce situation, the husband can leave whenever he wants; he isn’t obligated to stay. However, women must seek ‘Khul’ through the court.” (MW-05, Amanat al-Asimah, urban, aged 35 or over)*

Divorced women had no rights or entitlements even if they invested financially in the household through their work.

> *“If a woman is married to a man, even for 20 years, and the husband divorces her, she does not get anything, unlike in other cultures where women receive a share of everything.” (MW-07, Amanat al-Asimah, urban, age 35 or over)*

Midwives reported that, during pregnancy, women often experienced insufficient access to food, psychological support, and healthcare services, resulting in a higher risk of malnutrition and postpartum depression.

> *“The pressures on the family have increased. Due to frequent childbearing, pregnant women don’t get proper nutrition or sufficient psychological care. This is violence in itself, I believe.” (MW-06, Amanat al-Asimah, urban, age 35 or over)*

Women were often confined to domestic duties, while men received prioritised access to education and healthcare services. Midwives reported that girls are more likely to discontinue their education compared to boys. Midwives primarily attributed this trend to the undervaluing of girls’ education, the scarcity of girls’ schools, and parents’ concerns about their daughters’ safety during travel to and from school.

> *“In society, there’s a common belief that women will eventually marry. There’s a preference for educating male children because they carry on the family name.” (MW-09, Sana’a Gov, rural, age 25-34)*

Due to the preference for male children, midwives said that women who did not produce a male child may face divorce.

> *“I witnessed a case where a husband said to his wife during childbirth, ‘If you do not give birth to a male child, I will divorce you.’ He then divorced her.” (MW-10, Sana’a Gov, rural, age 35 or over)*

Another midwife described some situations where delivering a baby girl creates problems between couples or leads to separation.

> *“Preferring a male baby over a female is a type of violence we face during our work. The husband can cause problems during childbirth. Our job is to raise awareness in these situations. My colleagues in family planning have seen similar cases.” (MW-18, Aden, urban, age 35 or over)*

### Economic violence

Midwives mentioned several forms of economic violence, including failure to provide for the family’s basic needs, which often leads to disputes among the couple, which are exacerbated if the woman is employed and the husband depends on her to support the family.

> *“Violence begins when men fail to meet the basic needs of women. Another form of financial violence occurs when a woman relies on her husband financially, yet he withholds essential requirements” (MW-02, Amanat al-Asimah, urban, age 25-34)*

> *“Economic violence happens to the employed woman; for example, when the man sees that the woman is employed, he neglects her rights, even if he is financially capable. I am talking here from personal experience.” (MW-15, Taiz, urban, age 35 or over)*

Midwives found that unmarried daughters bore substantial responsibilities for supporting their parents and siblings. Families often viewed employed daughters as their only source of income and may prevent them from marrying to retain their financial support.

> *“I know a midwife working at a clinic in a remote area. She was working with a colleague of hers, an assistant doctor. He proposed, but her family refused him because they feared losing her financial assistance.” (MW-09, Sana’a Gov, rural, age 25-34)*

When women file for divorce, midwives report that their husbands will take full custody of the children and may take control of their ex-wife’s monetary and non-monetary assets. Midwives found that husbands forced their wives into selling their gold or giving them money to spend on non-essentials.

> *“When a husband fails to pay alimony, it is considered a form of violence. I personally know a lawyer who supports her children financially because she was unable to obtain child support from her former husband, as this process involves a lengthy legal procedure.” (MW-07, Amanat al-Asimah, urban, age 35 or over)*

### Sexual and reproductive health and rights-related violence

Midwives felt that not allowing women and girls to exercise their SRHR was an important form of VAWG.

> *“Violence against women includes underage marriage, early childbirth, continuous childbearing preventing women from taking birth control pills, not taking good care of her during pregnancy or giving her the rights of pregnant women.” (MW-07, Amanat al-Asimah, urban, age 35 or over)*

Midwives reported that women and girls frequently cannot decide whether to have a facility- or home-based delivery. Often, the husband or mother-in-law will decide, particularly in rural areas where home deliveries are more customary. Husbands may not allow their wives to have a facility-based delivery without being guaranteed that only female providers will deliver the child.

> *“Yes, most likely, this decision is made by the man or her mother-in-law. They usually say, “We all delivered at home. Why don’t you deliver at your house like we did? Why do you want to go to the hospital? We can provide you with a Jeddah or a midwife.” (MW-20, Aden, urban, age 35 or over)*

Similarly, midwives reported that women and girls need their husband’s permission to access long-term birth control methods like implants or injections, which limits their access to family planning.

> *“As a midwife, I do not accept giving a woman family planning pills without the permission of her husband, as I am keen to avoid any problems…In a Yemeni society, the man is consulted, and his opinion is taken on all matters.” (MW-10, Sana’a Gov, rural, age 35 or over)*

If a husband discovers that his wife is using birth control without his permission, she may face divorce or physical violence. Not only do women face threats, midwives also encounter verbal and physical violence when providing women with contraceptives.

> *“A woman with three children came to us for birth control. After counselling, she chose to take the pills. I suggested, ‘Why don’t you get an implant like your brother’s wife?’ She asked if her brother’s wife had visited. I realised she had come secretly. The next day, her husband came and accused us of cutting reproduction [performing sterilisation]. I was young and made a mistake; my tongue slipped, but I’ve learned from it. Now, I work in a different centre in Sana’a where people have a better mentality than in Khamer.” (MW-09, Sana’a Gov, rural, age 25-34)*

Midwives also reported significant reproductive consequences of violence, including unintended or underage pregnancy, pregnancy loss, and obstetric fistula.

> *“My sister-in-law suffered abuse from her husband, causing her to miscarry during pregnancy. He even took her gold, and after losing everything, she asked for a divorce.” (MW-08, Sana’a Gov, rural midwife, age 35 or over)*

> *“Cases with obstetric fistula continue to increase; in the past, they were rare. I believe that the main reasons are obstructed labour resulting from the long distance to reach the nearest health service, non-functionality of the health facility, and the increase in early marriage.” (MW-01, Amanat al-Asimah, urban, age 35 or over)*

### Community violence

Midwives also described community violence, in which women encounter aggression from strangers in public areas. This included rape, abduction, and robbery. Midwives reported women also experienced violence in the workplace perpetrated by supervisors or co-workers.

> *“I saw a man pull a bag from a woman until he dislocated her shoulder; he was trying to kidnap her.” (MW-03, Amanat al-Asimah, urban, age 35 or over*

> *“There is a case where a woman was kidnapped by individuals driving a bus and subsequently raped. A similar incident occurred with a woman I know; she was on her way home at night, boarded a bus, and was kidnapped by the bus driver, who then raped her. Despite resisting and defending herself, she sustained numerous injuries and sought assistance from us.” (MW-16, Aden, urban age 35 or over)*

### Violence against women in rural versus urban areas

Midwives highlighted differences in VAWG between women and girls in urban and rural areas. In rural settings, women and girls were responsible for agricultural labour, fetching water, and caring for their households and children.

> *“A village girl performs much harder work compared to a city girl. For example, she fetches water from distant, often mountainous areas and takes care of housework. If she doesn’t complete her tasks, she may face violence from her husband.” (MW-13, Taiz, urban, age 35 or over)*

Pregnant women were exempted from agricultural work, which heightened the risk of miscarriage and exposure to pesticides.

> *“Violence can be from the parents or a husband. Physical violence is not limited to beatings, but also, through assigning women hard work such as farming and using pesticides during pregnancy.” (MW-7, Amanat al-Asimah, urban, age 35 or over)*

### Conflict and pandemic-related violence

Midwives emphasised that in situations of displacement and insufficient support, women faced an increased risk of exploitation and abuse. They highlighted instances where women who had lost their husbands in the war were pressured to remarry, only to end up being treated as domestic servants by their new spouses.

> *“Violence has increased after the war due to poverty, affecting the mental health of husbands and wives, leading to conflicts between them. When husbands leave for the war front, their absence leaves the family without a provider, placing heavy responsibilities on the wife. This has led to a rise in widows who sometimes marry again and work as maids, exposing them to abuse. Girls are also affected, being deprived of education and forced to undertake tasks like fetching water and firewood.” (MW-06, Amanat al-Asimah, urban, age 35 or over)*

Midwives indicated that COVID-19-related restrictions to movement were related to an increased risk of IPV. Women were burdened with additional responsibilities during the pandemic, further increasing the gender-inequitable distribution of household responsibilities.

> *“When my husband got COVID-19, it really changed him. Men cannot bear illness. With our low income, I had to take on extra work at a clinic to cover his treatment costs, which were around 10,000 Riyal per day. The pandemic put a lot of strain on us financially, and surely many other women faced similar pressures.” (MW-08, Sana’a Gov, rural, age 35 or over)*

### Root causes of violence against women

Midwives saw deep-seated gender inequalities, traditions, and norms, and the war and pandemic-related economic crisis as critical drivers of VAWG in Yemen. Most Yemeni adults of working age were either unemployed, underemployed or, as for public sector workers, no longer paid due to the war and pandemic. Northern regions suffered most from economic hardship; southern regions were most affected by severe inflation.

> *“Patriarchal society is a reason for violence. A father abuses his daughter through early marriage, and a husband mistreats his family due to poor living conditions. Job opportunities have decreased, rental prices have risen, and prices in general are high and because of the war wages have been cut. Women have become a burden on men.” (MW-01, Amanat al-Asimah, urban, aged 35 or over) “Especially as I mentioned earlier, the ongoing challenges of war and poverty in our country have worsened our economic situation and contributed to a rise in domestic violence. We suffer from violence due to the economic situation and the psychological problems faced by men.” (MW-04, Amanat al-Asimah, urban, age 35 or over)*

> *“The economic conditions greatly worsened domestic violence, in addition to the customs, traditions, and society, especially in the rural areas.” (MW-01, Amanat al-Asimah, urban, age 35 or over)*

Midwives cited gender-inequitable laws and norms as an essential barrier to addressing VAWG in Yemen. Midwives highlighted the impact of tribal law as a source for perpetuating certain types of VAWG, including early marriage.

> *“As I mentioned earlier, in a tribal society, it’s very challenging for women to report violence, especially if it’s domestic violence.” (MW-04, Amanat al-Asimah, urban, age 35 or over)*

> *”Due to our customs and traditions as a tribal community, the legal system is almost non-existent, and tribal leaders often settle disputes based on customary practices. There are no prohibitions within these customs and traditions against marrying girls at the age of twelve.” (MW-04, Amanat al-Asimah, urban, age 35 or over)*

Daughters were generally considered possessions of their fathers, giving the fathers the authority to arrange marriages or exchange their daughters for financial gain. This is in addition to the deep-rooted belief that marriage protects’ women and girls’ and consequently the family’s honour.

> *“The customs of Yemeni society and their belief that marriage is a cover for women.” (MW-10, Sana’a Gov, rural, age 35 or over)*

### Intersectionality and VAWG

Midwives reported that women and girls from impoverished, marginalised backgrounds were more vulnerable to interpersonal violence due to a combination of discrimination, poverty, low socioeconomic status, and limited access to food, education, and healthcare services, including family planning. Midwives reported that the most vulnerable women and girls from marginalised groups were more likely to experience sexual violence and early marriage. For example, women and girls who do not have male relatives were at higher risk for sexual violence.

> *“Men may become more abusive when there are no relatives, brothers, or when a girl is an orphan or abandoned by her family.” (MW-01, Amanat al-Asimah, urban, age 35 or over)*

> *“The marginalised are more vulnerable to underage marriage, early pregnancy, childbirth problems, anaemia, and lack of hygiene.” (MW-12, Taiz, urban, age 35 or over)*

Midwives who worked in Taiz suggested that VAWG was more prevalent among a local ethnic minority group, *Al-Mohamasheen*, that faces high levels of discrimination in Yemen. Midwives felt that

*Mohamasheen* women and girls were disproportionately affected by IPV and societal GBV, specifically early and very early marriage, child labour, and a lack of access to formal education. Midwives reported that *Mohamasheen* parents had to force their daughters to drop out of school and engage in begging, street work, or early marriage because of the family’s inability to cover their basic economic needs.

> *“Many women experience verbal and physical abuse, particularly among marginalised and impoverished groups, such as the Al-Mohamasheen. Because they are very poor, they might be tempted by money and subsequently become victims of sexual exploitation.” (MW-20, Aden, urban, age 35 or over)*

> *“…there is a deprivation of girls from education. Many girls drop out of school and are forced to work. Those who do not work are often required to fetch water from the well. In addition, families that own land often require their daughters to work in the fields. For example, girls from marginalized communities typically do not continue their education beyond the fourth grade before they are made to work on the land.” (MW-12, Taiz, rural, age 35 or over)*

> *“It is common to find an 11-year-old girl from a marginalized group who is married. When I asked her why she married at such a young age, she replied, ‘We don’t have enough rooms in the house.’” (MW-12, Taiz, urban, age 35 or over)*

### Reporting violence

Midwives reported several barriers that prevent victims of violence, particularly sexual violence, from reporting that violence or otherwise seeking help.

> *“If a girl runs away and goes to another secure place, this ruins her reputation; even if she is a victim, people’s opinion is that her father has the right to discipline her.” (MW-14, Taiz, rural, age 25-34)*

Midwives said that a woman who reports or manages to escape violence could experience greater violence as retribution from the abuser or be labelled as “deviant” or “uncultured” and ostracised by her family and community.

> *“A woman who reports domestic violence would be rejected by society, labelled as rebellious, tomboyish, and ostracised by her family.” (MW-02, Amanat al-Asimah, urban, age 25-34)*

In other cases, midwives said that women who reported violence were not believed.

> *“If a woman goes to complain about her husband, society assumes she is lying, even without knowing the details of what happened.” (MW-15, Taiz, urban, age 25-34)*

Midwives said that women and girls who experience or report sexual violence are often blamed for “enticing” the perpetrators.

> *“The community believes that if a girl is respectful, she will not face any harassment, and no one will dare to assault her. However, if she’s not seen as respectful, she’s thought to deserve many immoral comments.” (MW-16, Aden, urban, age 35 or over)*

Midwives reported that some women or children who leave violent situations or report are at a higher risk of experiencing more violence.

> *“The violence escalates further, especially if the girl leaves her home and goes to another place. This tarnishes her reputation, even if she is a victim of violence. People believe that her father is the one who disciplined her. This violates women’s rights and exposes them to more violence.” (MW-14, Taiz, rural, age 25-34)*

> *“In our society, if a girl commits suicide, it might be covered up as a car accident or something else. Parents won’t admit that abuse at home was the reason for her suicide.” (MW-01, Amanat al-Asimah, urban, age 35 or over)*

Midwives reported that sexual violence committed within the family, such as by a father or brother, is highly stigmatised. Survivors of familial sexual violence find it very challenging to disclose their situation or seek help because of the high levels of stigma.

> *“There may also be relatives who commit sexual violence against family members. Girls are more susceptible to it, as they stay at home. If the girl speaks, she will be beaten and blamed.” (MW-1, Amanat al-Asimah, urban, age 35 or over)*

> *“Most cases who call the psychological support hotline are victims of violence, but they don’t disclose it. They call just to receive psychological support, also they don’t say that the problem is theirs. They say, a friend of mine, a neighbour I know.” (MW-01, Amanat al-Asimah, urban, age 35 or over)*

> *“If a woman experiences violence, she often avoids seeking help at hospitals due to the inhumane treatment she may receive. Consequently, women frequently choose to remain silent rather than endure further humiliation.” (MW-07, Amanat al-Asimah, urban, age 35 over)*

Women may opt not to report violence perpetrated not only by husbands but also by their older children out of fear of bringing shame or dishonour to the family.

> *“A mother came to the centre with bruises on her body from the abuse by her children. I advised her to report it to the police, but she refused, fearing for her children’s reputation.” (MW-07, Amanat al-Asimah, urban, age 35 or over)*

Custody concerns further limit women and girls’ options if they do leave an abusive partner or family.

> *“In our village, when a woman divorces, they usually take her children away from her. However, based on my experience, I managed to get custody of my children from the court, even though their father was wealthy. This encouraged many women, and now many children are under the custody of their mothers or grandmothers.” (MW-07, Amanat al-Asimah, urban, age 35 or over)*

### The challenge of understanding and addressing violence

Midwives said that Yemeni society strongly opposes the concept of interpersonal violence.

> *“The term ‘violence’ itself is surrounded with many issues. Even within the Ministry [of Health] and its programs, there is a problem with the term ‘violence against women’ because our religion does not allow violence against women. People hide what happens to them, like when a husband insults his wife.” (MW-01, Amanat al-Asimah, urban, aged 35 and over)*

As such, midwives found that projects addressing interpersonal violence were met with substantial criticism from the government, community leaders, and the families the projects intend to serve.

> *“Any course related to gender-based violence requires extensive procedures within the Ministry of Health for implementation because society does not accept these things.” (MW-02, Amanat al-Asimah, urban, age 25-34)*

While sexual violence was generally seen as the victim’s fault and could lead to the survivor’s social isolation, imprisonment, or death, physical violence was perceived as a crime against women and children. Midwives reported that an abuser who physically harmed women could face intense social pressure from society, especially in tribal communities. Midwives said that, in cases of physical violence, the local Sheikh generally holds the abuser accountable, and the abuser may be socially isolated.

> *“Society does not accept violence against women; even a man who abuses his wife or his children falls from society’s eyes, so violence is usually hidden and rarely reported by women.” (MW-01, Amanat al-Asimah, urban, age 35 or over)*

> *“Physical violence more stigmatised [for the abuser] than other forms of violence.” (MW-11, Taiz, rural, age 35 or over)*

Midwives found that survivors of sexual violence were not treated respectfully in health facilities, which prevented them from accessing support in the case of sexual violence and violence-associated pregnancies.

> *“If a woman experiences violence, she often avoids seeking help at hospitals due to the inhumane treatment she may receive. Consequently, women frequently choose to remain silent rather than endure further humiliation.” (MW-07, Amanat al-Asimah, urban, age 35 or over)*

### Survivor’s response and positive deviance

Given the barriers to and consequences for reporting violence, midwives reported that most women and girls remain with their abusive husbands to preserve their marriage and ensure the well-being of their children.

> *“When a woman experiences violence, she often remains silent because when she turns to her family, they usually send her back to her husband’s home, exposing her to more violence. Therefore, she stays silent most of the time.” (MW-08, Sana’a Gov, rural, age 35 or over)*

Midwives said that, in extreme cases, women or girls might turn to a friend or their family of origin for protection.

> *“…her friend was the first option [when she left her abusive husband], followed by her relatives. I know a friend of mine who experienced violence and then went to live with her friend.” (MW-08, Sana’a Gov, rural, age 35 or over)*

A few participants mentioned that women or girls may turn to their children for support in mitigating or preventing violence.

> *A woman who experiences violence turns to her family. If she has children, she uses them to protect herself, which causes problems between the father and the children.” (MW-12, Taiz, rural, age 35 or over)*

Women or girls who were raped could sometimes access birth control or emergency contraception provided by midwives to prevent pregnancy.

> *“Women who experience sexual violence often choose to remain silent and endure the situation, or they confide in a friend. In certain instances, they resort to taking birth control pills, particularly if they have been assaulted by family members such as brothers or fathers. I know a specific case where a father raped his daughter.” (MW-03, Amanat al-Asimah, urban, aged 35 or over)*

Sometimes, women or girls also turned to their in-laws for support. When no support was available from within their family network, women and girls might seek assistance outside the family, especially when they feared for their lives.

> *“When a woman feels unsafe at home and cannot bear violence any longer, she may take legal action. Initially, she often seeks intervention from her husband’s family, discussing the matter with his brothers and mother. If her concerns are ignored, her last option may be to seek legal separation through the courts, especially if the violence worsens.” (MW-17, Aden, urban, age 35 or over)*

Despite the protracted and costly legal procedure, midwives reported that women sometimes pursued divorce or attempted to retain custody of or receive alimony for their children.

> *“I am a survivor of domestic violence. I was abused by my husband, and after three years of legal proceedings, I divorced him with strong evidence. I was granted a monthly alimony of six thousand for my child, but it has not been enforced yet.” (MW-07, Amanat al-Asimah, urban, age 35 or over)*

That said, most participants indicated that women and children would rarely go to the police to report family violence due to fear of escalation of violence or of losing custody of their children.

> *“During my work, if a case of rape or physical violence is identified, she chooses whether she wants to report her incidence or not, and most of the cases think about their children, so they prefer not to speak.” (MW-02, Amanat al-Asimah, urban, age 25-34)*

Midwives reported that the local tribe’s Sheikh and community were essential resources for women and girls who experienced physical violence.

> *“In a village far from our village, a woman went to the Sheikh because her husband beat her and locked her in the bathroom. She screamed out of the bathroom window, and people gathered, brought the Sheikh of the neighbourhood and went to help her.” (MW-11, Taiz, rural, age 35 or over)*

> *“There is a married woman in another village whose husband beats her, so she takes refuge at night in the Sheikh’s house until the morning when her family comes to her.” (MW-06, Amanat al-Asimah, rural, age 35 or over)*

Midwives did not report instances where Sheikhs assisted women and children who experienced violence other than physical violence. Midwives suggested that survivors’ responses to violence were influenced by various factors, including their background, education, family support, and financial independence. Women who were employed and had a support system in place were more likely to take decisive actions such as reporting the violence, pursuing divorce through procedures like *Khul,* a procedure that allows women to divorce their husbands, and seeking custody of their children through legal means.

> *“Not all women choose to report violence; instead, some endure it, particularly if it’s limited to verbal abuse. Certain women tolerate difficult living conditions and verbal mistreatment for the sake of their children. However, financially independent female employees opt for divorce, resorting to Khul.” (MW-04, Amanat al-Asimah, urban, age 35 or over)*

In contrast, midwives highlighted that women who were financially dependent on their husbands often felt powerless and had to tolerate the violence they experienced silently.

> *“Many abused women choose to remain patient and silent, while educated, financially independent women may seek assistance from the courts.” MW-07, Amanat al-Asimah, urban, age 35 or over)*

Beyond resorting to the local Sheikh or their family of origin when violence became intolerable or turning to midwives to prevent sexual violence-related pregnancies, midwives did not describe women or girls accessing resources outside of their communities (e.g., domestic violence shelter) for support when confronting interpersonal violence.

> *“Yes, there is a woman whose husband beat her and divorced her. The woman resorted to the neighbourhood Sheikh, who referred her to me to check if she was abused, so the Sheikh took her for a day and ordered the husband to leave the house for her and her children.” (MW-08, Sana’a Gov, rural, age 35 or over)*

> *“A woman who experiences violence may be unaware of the places available for her protection, or she may fear her husband, worry about leaving her children, or find it difficult to access these resources.” (MW 05, Amanat al-Asimah, urban, age 35 or over)*

### Family and community support for women and girls who experience violence

Midwives found that the level of family support for women facing violence varied depending on the type of violence and the socioeconomic status of both the family and the perpetrators.

> *“When she returned to her family, they beat her and sent her back to her husband because he was wealthy.” (MW-01, Amanat al-Asimah, urban midwife, age 35 or over)*

Families and communities tended to offer more support to women or girls who experience physical violence than emotional, economic, or sexual violence.

> *“Her family always advises her to remain patient, but if the abuse escalates to physical violence, it is not tolerable.” (MW-01, Amanat al-Asimah, urban, age 35 or over)*

Families with the means provided their daughters with financial and emotional support when they experienced physical violence from their husbands.

> *“If they have the financial means, they will support her. If they cannot help her, they will temporarily host her and then return her to her husband’s house.” (MW-05, Amanat al-Asimah, urban, age 35 or over)*

Sometimes, supportive families were not able to accept the women or girls with their children into their homes due to the additional financial burden and the belief that the father, rather than the mother, is responsible for child-rearing. Not all families were willing are able to take their daughters in.

> *“If her family is supportive, they protect her, but if they are not supportive, they send her back to her husband’s house. Then she must endure violence.” (MW-05, Amanat al-Asimah, urban, aged 35 or over)*

> *“Some families take their daughter, some provide financial support, and some families do not believe their daughters.” (MW-03, Amanat al-Asimah, urban, age 35 or over)*

## Discussion

In this cross-sectional, qualitative study, 20 NYMA midwives reported on the root causes, forms, perpetrators, and consequences of VAWG and how women and girls tried to respond to, prevent, and mitigate the violence that they faced. In keeping with global research on VAWG, midwives highlighted IPV as the most common form of VAWG [1], but also described violence from women and girls’ own families and mothers-in-law seeking to protect a woman or girl’s “honour.” Midwives described how VAWG resulted in a spectrum of consequences, including unwanted pregnancies, multiple childbirths, unsafe abortions or deliveries, pregnancy loss, excessive bleeding, obstetric fistulas, and mental health issues, including depression and suicide. We summarise challenges to and mitigation strategies for addressing VAWG in Yemen in Figure 1.

**Figure 1.** Different levels of VAWG in Yemen, their root causes, and mitigation strategies as reported by community midwives

Midwives discussed how the conflict, displacement, and protracted economic crisis have heightened the risk of early and forced marriage and compounded barriers to education, employment, health care, and food, in keeping with other studies suggesting that IPV is more pervasive in crises [23]. Given the lack of security and the centrality of women and girls’ “honour” to family honour, families may see marrying off their daughters as the only or best way to protect them from rape [24]. These findings are consistent with a UN report that indicated an increase in VAWG, including early marriage, in response to the war and COVID-19 in Yemen [25], and a study conducted by UNFPA Women’s Refugees Commission that identified a rise in early child marriage among the displaced population as compared to the host community in three Yemeni governorates [7]. Midwives’ report that child marriage and other forms of VAWG have increased in response to the ongoing conflict and that gender-inequitable cultural norms contribute to VAWG align with the findings from studies conducted in similar conflict-affected settings in South Sudan and Palestine [26,27], and with those of a recent systematic review of the prevalence of sexual violence among female refugees and internally displaced people in 14 countries which found that 21% had experienced sexual violence [28].

In addition to the ongoing conflict, midwives felt that patriarchal norms and gender inequitable laws and policies were a significant cause of VAWG that also prevented women and girls from responding to VAWG beyond tolerating the violence. This finding is in keeping with research related to how gender inequitable norms and limitations on the freedom of women and girls [29], the loss of economic, family, and social capital, and the collapse of institutions for governance, security, and justice [27] relate to increased VAWG. Emerging research on the relation between conflict and patriarchal norms suggests that gender disparities and patriarchal attitudes also become pervasive in conflict and crises [30,31] further highlighting the need to develop local approaches to addressing gender inequitable norms.

Midwives explained how the most marginalised women and girls are at the highest risk of exploitation and abuse, which is supported by studies on intersectionality and VAWG [32]. Midwives identified that marginalised, impoverished, orphaned, and ethnic minority women and girls were particularly susceptible to violence. They also found that women residing in rural areas faced additional forms of violence, including physically demanding agricultural work while pregnant and exposure to harmful pesticides. Midwives reported that women and girls in rural areas had more limited access to education and were more likely to experience child marriage than women and girls in urban areas.

Midwives highlighted several strategies women and girls use to prevent, mitigate, or escape violence, all of which involved resources available to them within their communities. The first and often the only strategy was to tolerate the violence. The local Sheikh was seen as a trusted resource for women and was sometimes asked to mediate between the husband and wife or their families in the context of physical violence. Midwives said that women and girls only sought help from their families or their local Sheikh when the physical violence became intolerable or they feared for their or their children’s lives. While midwives said that women and girls might turn to their local Sheikh in cases of extreme physical violence, no midwives described women and girls going outside of their local community for help to access a domestic violence shelter. Midwives reported that the level of family support for women and girls facing violence varies depending on the type of violence and the socioeconomic status of both the family and the perpetrator’s family. Midwives shared incidents where women who were subjected to beatings were sent back to their husbands because their husbands were well off financially and better able to support them and their children than their families of origin. Families and communities tended to offer more support to women experiencing physical violence, such as beatings, than for emotional or economic violence, which women were expected to tolerate, and sexual violence, which was highly stigmatised.

Midwives described how they supported women and girls who experienced sexual violence through the provision of birth control pills or emergency contraception but did not mention any other examples of women and girls seeking help for sexual violence. For midwives, offering contraceptive methods could itself be dangerous, and midwives described situations where women need to carefully consult with or obtain permission from their husbands before seeking contraceptives. As described by midwives, sexual violence is highly stigmatised, and women or girls who experience sexual violence can be themselves jailed or killed. In addition to intense social pressures that prevent reporting and other help-seeking behaviours, other studies suggest that economic barriers moderate access to the limited resources available for supporting women and girls who experience violence in crisis [25]. While there is a domestic violence shelter in the capital city, Sana’a, other services that may have been available before the onset of the war in 2015 may have been suspended. No midwives mentioned women or girls who experienced violence as seeking help from the one domestic violence centre that we identified, which suggests that women and girls may not be aware of the shelter or that social, cultural, economic, and societal- and conflict-related security concerns may prevent women and girls who experience violence from accessing the only existing shelter. Resources and support services addressing VAWG could be disseminated more widely through social media, hotlines, radio, and TV to build awareness, although lack of awareness is likely one of the many barriers affecting women’s access to the shelter.

Midwives discussed important difficulties in providing training or other support for midwives to better serve women and girls who experience violence. For example, they said that the concept of physical violence against women and girls, generally referred to as beatings, was highly stigmatised for perpetrators and that sexual violence was highly stigmatised for survivors. They also mentioned that trainings and projects for health workers that mentioned the term “violence” might be met with resistance from local or national government agencies responsible for approving or overseeing these trainings or interventions.

These findings underscore the importance of local approaches to supporting women and girls who experience violence and of human design or positive deviance-centred approaches to safety planning where women and girls who experience violence access local resources to safely mitigate, prevent or otherwise address the violence that they experience. Both funders and groups supporting maternal and child health (MCH) in conflict-affected settings must understand that violence-related training and interventions cannot be developed without these formative studies to clarify the local context, values, preferences, and constraints. The lack of funding to support formative studies, like this one, and the prework needed to ensure that VAWG research and interventions are safe for the research team, service providers, and women and girls, and in keeping with local laws, values, and preferences contrasts with the clear need to invest in this work to address the deeply entrenched and prevalent challenge of VAWG. Maintaining confidentiality, providing quality training to those collecting data, ensuring research participant and interview safety, and providing support services for research participants and interviewers are central to the ethical conduct of VAWG research [19]. These issues often cannot be adequately addressed within the limited budgets allotted to VAWG research, especially given the limited funding for VAWG research within conflict-affected and displaced populations, like women and girls in Yemen.

### Next steps for supporting women and girls who experience violence in Yemen

Given the ongoing conflict, Yemeni women and girls are in urgent need of health and protective services, including basic needs, medical and legal assistance, and mental health support [25,33]. Given the high levels of stigma associated with VAWG, services for survivors should be integrated into the existing healthcare system rather than offered as stand-alone services [34]. In Yemen, where 80% of the population lives in rural areas, the war has decimated the formal health and education systems, and women and girls have limited mobility and may not be able to access the formal health system without a male relative, community midwives may represent the only intersection between women and girls who experience violence and the formal health system. Midwives could be trained to screen women and girls for violence-related complications during routine health services and to provide them with information about available support services within their communities. Incorporating violence-related healthcare into routine healthcare can help improve women and girls’ ability to use these services as it removes the stigma of accessing the services and may encourage women and girls to report the violence they have experienced and seek help from a trusted community research, like community midwives, albeit in a way that is safe and acceptable to them [34].

Midwives and other trusted healthcare providers should be able to support women and girls experiencing violence without fearing for their own safety. International donors should fund the local development and implementation of initiatives that educate the public about VAWG, support economic development to reduce the drivers of VAWG, and empower women, and encourage their education. Intervention programs, like incentives for parents to send their daughters to school or teaching women skills to help them obtain financial independence must consider local cultural norms and potential adverse consequences of women’s empowerment programmes. Further research is essential to inform and develop interventions against VAWG and to ensure decision-makers and policymakers prioritise addressing VAWG in Yemen. While several studies reported that the incidence and severity of VAWG increased in displaced communities and during humanitarian crises in Yemen, no studies have addressed the prevention of VAWG in Yemen [5,15]. At the global level, few studies have evaluated VAWG prevention programs targeting married, out-of-school, and displaced communities, which are essential concerns in the Yemeni context [35].

Yemen’s legal system does not set a minimum marriage age or give equal access to divorce, inheritance, child custody, or legal protection for men and women, which places women and girls at higher risk for domestic and sexual violence and reduces their options for addressing violence [9,13]. Midwives’ reports that gender inequitable laws restricted women and girls’ options underscore the need for a locally developed policy agenda for addressing VAWG. Community and school-based education campaigns could support changes to gender-inequitable cultural norms around VAWG.

In the absence of funding, whether or not to conduct this research without the means to offer services or support to the research team and the midwives who shared their experiences working with women and girls who experience violence was an important ethical question. Before conducting this work, LM had written four grants to fund this formative research, all of which had very low odds of success (2-5%) for grants with very restricted budgets, which would have been inadequate for conducting the research and providing support services for the research team and participants. None of the grant applications was funded, meaning the fieldwork had to be self-funded by the PI. One grant application for 100K for formative research to a funder of research in humanitarian settings was rejected in part because, “researchers did not intervene to address VAWG.” These types of expectations for formative research on the context where VAWG happens dangerously disregard the importance of understanding the context before engaging in VAWG-related intervention research. While midwives were reporting on violence that they heard about or saw rather than their own experiences, witnessing and learning about violence is itself traumatic, especially as the options for women and girls who experience violence are so limited and mental health services should have been offered to participants. It is concerning that the global funding community has not made the necessary investments in VAWG-related research to address the scale of the problem adequately.

### Strengths and limitations of this study

To our knowledge, this study represents the first attempt to engage midwives in VAWG-related research in Yemen. While limited in scope, the findings from this formative research provide a basis for further research on VAWG in Yemen and highlight the role of midwives as a group who closely interact with women and girls and confront manifestations of VAWG in their daily work, often while facing similar violence in their lives or as a result of helping those facing violence. This research had some limitations. Conducting VAWG research in a humanitarian crisis presents critical ethical and logistical challenges, including issues related to participants’ informed consent, confidentiality, and the safety of the interview team and participants, and increased difficulties in availability of or access to resources to support women and girls who experience violence [33]. While we asked midwives to describe their understanding of VAWG experienced by women and girls that they served, which can itself be traumatising, midwives sometimes reported on the VAWG they themselves experienced and they reported on violence they experienced in the course of their regular work, as in the context of providing family planning. Ideally, we should have developed safety plans and secured mental health services for the midwives who participated in this research even though they were not asked about their own experiences of violence. The protracted conflict, COVID-19 pandemic, and very restricted budget for this work meant we wanted to limit interviewer and participant travel. As such, we condensed our interview timeline to accommodate midwives’ regular travel, meaning we did not have time to transcribe, translate, and code transcripts as the research was conducted. To mitigate this limitation, the team discussed the interviews following their conduct to ensure the guide incorporated findings from past interviews. Due to the closure of Sana’a International Airport and security issues during the time interviews were conducted, some interviews were conducted by phone due to the difficulty and insecurity of accessing some regions in Taiz. Conducting VAWG-related research is very sensitive and limited by political agendas and decision-makers [7,13]. Violence against LGBTQI and other marginalized groups, including Somali and Ethiopian minority women, women and girls with disabilities, and against men and boys are important topics that were not well addressed in this formative research.

## Conclusion

The ongoing war in Yemen has set back meaningful advances in addressing child marriage and labour and reduced the mechanisms available for women and girls to avoid, mitigate, confront, or otherwise address the violence in their lives. This study provides essential information on the types of violence women and girls experience, the underlying causes of that violence, how survivors and communities respond to VAWG, and barriers to reporting or otherwise addressing violence. Midwives play a crucial role in supporting women and girls who experience violence in Yemen. NYMA is a long-standing national network and NYMA-trained midwives act as trusted members of their communities, representing an important resource for women and girls who experience violence. Providing training and support to community midwives so that they can safely serve women and girls who experience violence should be a global development priority. In addition to midwives, the local Sheikh was seen as a trusted resource for women. Future work could consider creating opportunities for midwives to work with local Sheikhs to support women and girls who experience violence in Yemen. Cultural norms and stigma prevent disclosing, reporting violence, or accessing the minimal support services that exist. Making funding available to support community-led work to understand and address VAWG in Yemen should be a priority for global funders who support work to understand and improve MCH, women and girls’ SRHR, and the related SDGs.

## Data Availability

De-identified interview transcripts are available for download through ICPSR. https://doi.org/10.3886/E213941V1

## Acknowledgements

The authors would like to thank the National Association of Yemeni Midwives for coordinating the interviews with midwives and hosting interviews conducted in Aden, as well as the midwives who provided their experiences and insights for this study. The authors would also like to thank Mirna Naccache for her thoughtful edits to the manuscript. MAZ would like to thank the Else Kröner-Fresenius-Stiftung within the Heidelberg Graduate School of Global Health for supporting the first year of her work on this project. She would also like to thank the University of Sana’a for facilitating the local ethical clearance for this study. The authors would also like to thank Alankreeta Bharali for her assistance in formatting Figure 1.

## Contributors

MAZ, LM conceived of the study and wrote the study protocol. MAZ conducted the interviews. MOAZ transcribed the interviews. MAZ, LL, and LM developed and refined the study codebook. MAZ, LL, and HX coded the interview transcripts and LM supervised the analysis. MAZ and LM wrote the first draft of the manuscript. MAZ, HZ, LL, RFP, PS, MNAZ, LB, AJ, and LM edited the final manuscript. All authors had full access to all the study data, take responsibility for data integrity and reliability of the analysis, and had final responsibility for the decision to submit for publication.

## Supporting information

S1. Table. Search strategy for literature review

S2 Table. Codebook

S3 Table. Original Arabic and English-language translations of participant quotes

S4 Table. COREQ (COnsolidated criteria for REporting Qualitative research) Checklist for reporting qualitative research

S1 Text. PLOS inclusivity in global health questionnaire

